# Shared and distinct pathways from anxiety disorder and depression to cardiovascular disease: a UK Biobank prospective cohort study

**DOI:** 10.1101/2025.02.14.25322279

**Authors:** Shinya Nakada, Carlos Celis-Morales, Jill P Pell, Frederick K Ho

## Abstract

**Aims:** Although anxiety and depression increase the risk of cardiovascular disease, no studies have examined their underlying mechanisms systematically, particularly contrasting the difference between the two of them. Our study aimed to examine the extent to which lifestyle, physical and metabolic factors mediate the associations of anxiety and depression with incident CVD.

**Methods:** A prospective cohort study was conducted using UK Biobank. Anxiety, depression, and incident CVD were ascertained through linkage to hospital and death records. A causal mediation analysis was performed for anxiety and depression separately examining multiple potential lifestyle, physical, and metabolic mediators. Cox proportional hazard models and log-linear models were used to derive indirect effect estimates and proportions mediated.

**Results:** A total of 254,695 participants were included. Both anxiety (HR 1.62, 95% CI 1.15–2.13) and depression (HR 2.15, 95% CI 1.75–2.64) were associated with CVD after adjusting for sociodemographic confounders. The strongest mediators were current smoking and higher waist-hip ratio which accounted for 12.2% and 12.8% of the excess risk from anxiety, and 17.2% and 13.8% from depression, respectively. The strongest metabolic mediators were systolic blood pressure for anxiety (10.2%) and CRP (10.7%) for depression. Systolic blood pressure was the weakest mediator for depression (4.0%).

**Conclusions:** Lifestyle and physical pathways to incident CVD may be common to both anxiety and depression, but shared metabolic pathways seem unlikely. Our findings inform which risk factors to target among people with anxiety or depression in order to reduce their higher risk of developing CVD.

**What is already known on this topic:**

- Anxiety disorder and depression are associated with an increased risk of cardiovascular disease.
- Some mechanisms were suggested for depression (e.g., obesity and inflammation) but there was no systematic investigation of pathways especially in comparing depression and anxiety disorder.

**What this study adds:**

- Our study showed that whilst smoking and central obesity were common mediators of both anxiety disorder and depression, systolic blood pressure was more specific to anxiety disorder and C-reactive protein to depression.

**How this study might affect research, practice or policy:**

- Our findings inform how best to target prevention strategies to reduce the higher risk of cardiovascular disease among people with established anxiety disorder or depression.

## Background

Anxiety disorder and depression often co-exist with cardiovascular disease (CVD). Studies reported that more than 30% of patients with CVD had anxiety disorder [1] and 15% had depression [2]. One explanation is that these mental health conditions may increase the risk of CVD. A meta-analysis found that people with anxiety disorder or depression had about 20% increased risk of incident CVD [3]. Since anxiety disorder and depression are common and increasing [4, 5], understanding the pathways to CVD could identify modifiable targets to reduce comorbidity.

Some previous studies have conducted mediation analyses. A cross-sectional study has demonstrated that lifestyle, inflammation, and metabolic syndrome mediated the association between distress and incident CVD [6]. A prospective Australian study has demonstrated that inflammation and obesity mediated the association between depression and non-fatal CVD [7] and a prospective USA study has reported that diabetes (odds ratio 1.04; 95% confidence interval [CI] 1.02–1.06) and hypertension (odds ratio 1.02; 95% CI 1.00–1.04) mediated the association between depression and CVD mortality [8].

However, these studies have limitations. Firstly, they did not systematically assess and compare a broad range of potential mediators including lifestyle, physical, and metabolic factors. Understanding the relative importance of these potential pathways is key to targeting interventions. Secondly, these studies have focused on depression to the exclusion of anxiety disorder. Because anxiety disorder may have a different aetiology, the findings of studies based on depression cannot be automatically applied to anxiety disorder.

Therefore, we aimed to examine the extent to which lifestyle, physical and metabolic factors mediate the associations between anxiety disorder and depression and incident CVD, contrasting the difference between the two of them.

## Methods

### Study design and participants

Our study is a prospective cohort study using data from UK Biobank. Between 2007 and 2010, over 500,000 participants aged 37 to 73 years were enrolled. Participants visited one of the 22 assessment centres across England, Scotland, and Wales to provide their information and biological samples. We excluded participants who experienced CVD before the baseline assessment, those who developed depression or anxiety disorder less than one year before or after the baseline assessment to reduce the risk of reverse causation between the exposures and mediators, and those who had missing data in covariates or mediators.

### Measurements

The exposures of interest were anxiety disorder and depression diagnosed at least one year before the baseline assessment and ascertained through record linkage to hospital admission data available from 2004: Health Episode Statistics (England and Wales) and Scottish Morbidity Records (Scotland). Anxiety disorder and depression were defined as an International Classification of Diseases, 10th revision (ICD-10) code of F40-43 and F32-33, respectively.

The outcome of interest was incident CVD (fatal or non-fatal) after baseline assessment ascertained from record linkage to hospital admission data and death certificates. Hospital admission data were available up to September 2021 in England, February 2018 in Wales, and July 2021 in Scotland. Death certificate data were obtained from the National Health Service Information Centre (England and Wales) and the National Health Service Central Register (Scotland) and were available up to September 2021 in England and Wales, and October 2021 in Scotland. Follow-up was censored at the date of hospitalisation or death, whichever occurred first. CVD was defined as an ICD-10 code of I20-25, I42.0, I42.6, I42.7, I42.9, I50, I60-64, or I110.

The potential mediators of interest were lifestyle factors (physical activity, TV viewing, diet quality, smoking status and sleep duration), physical factors (waist-hip ratio, body mass index [BMI], and grip strength), and metabolic factors (systolic blood pressure, haemoglobin A1c [HbA1c], low-density lipoprotein-cholesterol [LDL-c], and C-reactive protein [CRP]). Lifestyle factors were self-reported by participants using a touchscreen questionnaire at baseline. Physical activity was based on metabolic equivalent minutes (METs) per week derived from the validated International Physical Activity Questionnaire [9] and categorised into METs <1000 or ≥1000 per week [10]. TV viewing was categorised as <3 or ≥3 hours per day, based on the median. Diet quality was based on the cumulative dietary risk factors score, which has been reported previously [11]. In brief, participants were given 1 point for each of nine dietary recommendations met relating to: processed meat, red meat, total fish, milk, spread type, cereal intake, salt added to food, water, and fruits and vegetables. Therefore, the overall score ranged from 0 (least healthy) to 9 (most healthy) but was categorised as ≥4 or <4 based on the median value. Smoking status was categorised as current smoker or not (never or former smoker). Sleep duration was categorised as <7 or ≥7 hours per day, based on the median.

Physical factors were measured by trained staff at the baseline assessment and treated as binary variables. Waist-hip ratio was calculated as waist circumference/hip circumference, measured using a Wessex non-stretchable sprung tape and categorised as ≥0.90 or <0.90 for men and ≥0.85 or <0.85 for women [12]. BMI was calculated as weight/height^2^; height was measured by a Seca 202 stadiometer, and body weight was measured by a Tanita BC-418 body composition analyser. BMI was categorised as ≥30 kg/m^2^ or <30 kg/m^2^ [13]. Grip strength was measured using a Jamar J00105 hydraulic hand 101 dynamometer and categorised as <36.0 kg or ≥36.0kg for men and <21.0 kg or ≥21.0 kg for female, based on the median.

Metabolic factors were measured or samples were collected by trained staff at the baseline assessment and treated as binary variables. Systolic blood pressure was measured using an automated Omron device 0-255 and the use of anti-hypertensive medication was self-reported. Systolic blood pressure was categorised as ≥140mmHg or taking antihypertensive medication or not [14]. HbA1c was categorised as ≥48 mmol/mol or taking diabetic medication or not [15]. LDL-c was categorised as ≥4.9 mmol/L or taking cholesterol-lowering medication or not [16]. CRP was categorised as >3mg/L or ≤3mg/L [17]. All these factors were selected because they are risk factors for CVD and could be affected by anxiety disorders and depression.

Potential confounders were age, sex, ethnic group, and deprivation level. Age, sex, and ethnic group were self-reported by participants using the touchscreen questionnaire at baseline. Deprivation was based on tertiles of the Townsend area deprivation index, which was derived from the postcode of residence using aggregated data on unemployment, car and home ownership, and household overcrowding.

### Statistical analyses

The main analyses were conducted in three stages: a total effect was estimated; an outcome model and mediator models were built; and a causal mediation analysis was performed. In the outcome model, incident CVD was regressed on either anxiety disorder or depression, in a Cox proportional hazard model that included all of the potential mediators and confounders. In the mediator models, each one of the mediators was regressed, in separate log-linear models, on either anxiety disorder or depression including all potential confounders. In the causal mediation, after testing for interactions between anxiety disorder/depression and the mediators, total natural indirect effects (NIE) and proportions mediated were calculated using separate Cox proportional hazard models and log-linear models for each of the mediators, adjusted for all the confounders. The Cox proportional hazard models were used to estimate hazard ratios (HRs) and the log-linear models were used to estimate risk ratios. Standard errors and 95% confidence intervals (CIs) were derived from bootstrap procedures. Proportional hazard assumptions were checked by statistical tests based on Schoenfeld residuals. Participants with anxiety disorder were not excluded from analysis of depression and vice versa.

A sensitivity analysis was conducted to assess whether the additional use of self-reported anxiety disorder and depression at baseline affected the results. All analyses were conducted using R version 3.5.3 with a package CMAverse version 0.1.0.

### Ethics Statement

All procedures contributing to this work comply with the ethical standards of the relevant national and institutional committees on human experimentation and with the Helsinki Declaration of 1975, as revised in 2013. UK Biobank was approved by the North-West Multi-Centre Research Ethics Committee (Ref: 11/NW/0382). This work was conducted under the UK Biobank application number 7155.

### Consent Statement

Written informed consent was obtained from all individual participants included in the study.

### Patient and public involvement

No patients or the public were involved in setting the research question or the outcome measures, nor were they involved in developing plans for design or implementation of the study. No patients or the public were asked to advise on interpretation or writing up of results. The results of our research can be disseminated on the UK Biobank homepage.

## Results

Of more than 500,000 UK Biobank participants, 254,695 were eligible for inclusion in the main analyses (Figure 1). Participants with anxiety disorder were more likely to have unfavourable lifestyle, physical, and metabolic factors except for diet quality, which was worse than those without anxiety disorder (Table 1). Those with anxiety disorder were also more likely to develop CVD (5.8% vs 3.9%). Similarly, participants with depression were more likely to have unfavourable lifestyle, physical, and metabolic factors, apart from systolic blood pressure, and were more likely to develop CVD (7.3% vs 3.8%).

**Figure 1.**
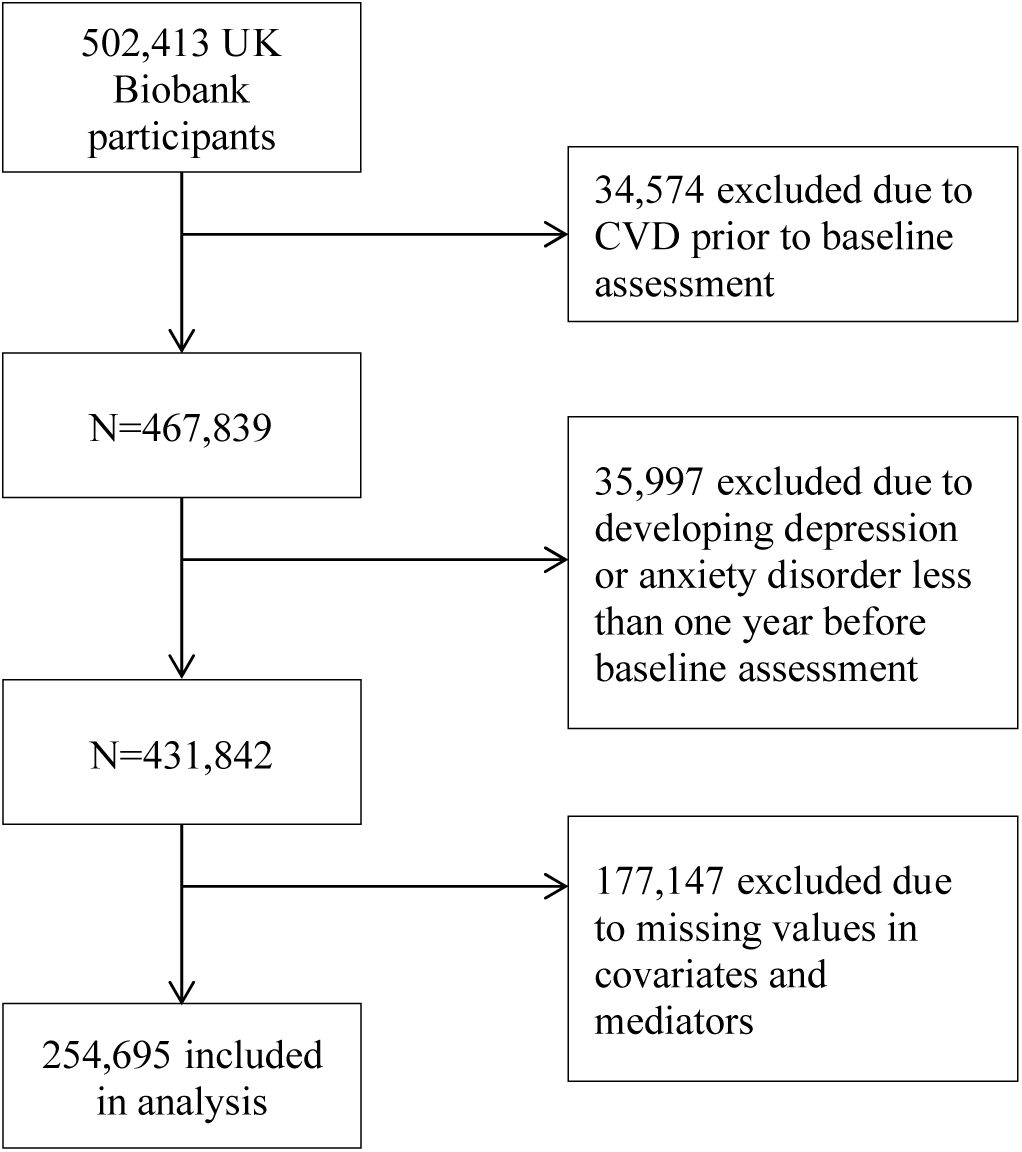
Flow chart of the participant selection process

**Table 1.** Characteristics of participants by anxiety disorder and depression.

|  | No anxiety disorder<br>N=254,145 (%) | Anxiety disorder<br>N=550 (%) | No depression<br>N=253,437 (%) | Depression<br>N=1,258 (%) |
| --- | --- | --- | --- | --- |
| Mean (SD) Age (years) | 55.8 (8.11) | 55.4 (8.25) | 55.8 (8.11) | 54.9 (8.01) |
| Sex: |  |  |  |  |
| Female | 135,788 (53.4) | 327 (59.5) | 135,363 (53.4) | 752 (59.8) |
| Male | 118,357 (46.6) | 223 (40.5) | 118,074 (46.6) | 506 (40.2) |
| Ethnic group: |  |  |  |  |
| White | 242,814 (95.5) | 525 (95.5) | 242,130 (95.5) | 1,209 (96.1) |
| Non-white | 11,331 (4.46) | 25 (4.55) | 11,307 (4.46) | 49 (3.90) |
| Deprivation level: |  |  |  |  |
| Least deprived | 88,546 (34.8) | 160 (29.1) | 88,402 (34.9) | 304 (24.2) |
| Middle | 86,027 (33.8) | 166 (30.2) | 85,854 (33.9) | 339 (26.9) |
| Most deprived | 79,572 (31.3) | 224 (40.7) | 79,181 (31.2) | 615 (48.9) |
| MET-minutes/week <1000 | 73,745 (29.0) | 180 (32.7) | 73,455 (29.0) | 470 (37.4) |
| TV viewing ≥3 hours/day | 122,642 (48.3) | 288 (52.4) | 122,213 (48.2) | 717 (57.0) |
| Diet quality score <5 | 72,556 (28.5) | 153 (27.8) | 72,284 (28.5) | 425 (33.8) |
| Current smoking | 22,931 (9.02) | 83 (15.1) | 22,732 (8.97) | 282 (22.4) |
| Sleep duration <7 hours/day | 58,865 (23.2) | 139 (25.3) | 58,624 (23.1) | 380 (30.2) |
| Waist-hip ratio |  |  |  |  |
| ≥0.90 (male) | 118,827 (46.8) | 297 (54.0) | 118,389 (46.7) | 735 (58.4) |
| ≥0.85 (female) |  |  |  |  |
| Body mass index ≥30kg/m<sup>2</sup> | 54,619 (21.5) | 147 (26.7) | 54,326 (21.4) | 440 (35.0) |
| Grip strength |  |  |  |  |
| <36 Kg (male) | 119,041 (46.8) | 298 (54.2) | 118,614 (46.8) | 725 (57.6) |
| <21 Kg (female) |  |  |  |  |
| SBP ≥140mmHg or medication | 120,996 (47.6) | 278 (50.5) | 120,682 (47.6) | 592 (47.1) |
| HbA1c ≥ 48 mmol/mol or medication | 7,845 (3.09) | 28 (5.09) | 7,802 (3.08) | 71 (5.64) |
| LDL-c ≥ 4.9 mmol/L or medication | 48,536 (19.1) | 131 (23.8) | 48,306 (19.1) | 361 (28.7) |
| CRP >3mg/L | 51,432 (20.2) | 144 (26.2) | 51,151 (20.2) | 425 (33.8) |
| 495 N, number; SD, standard deviation; MET, metabolic equivalent; TV, television; SBP, systolic blood |  |  |  |  |
| 496 pressure; HbA1c, haemoglobin A1c; LDL-c, low-density lipoprotein cholesterol; CRP, C-reactive |  |  |  |  |
| 497 protein |  |  |  |  |

Both anxiety disorder (HR 1.62, 95% CI 1.15–2.13) and depression (HR 2.15, 95% CI 1.75– 2.64) were associated with CVD after adjusting for sociodemographic confounders (Supplementary Figure 1). Anxiety disorder was associated with all potential mediators except for diet quality score and sleep duration and these mediators were associated with CVD. Similarly, depression was associated with all mediators but hypertension (Supplementary Tables 1 and 2).

Of the lifestyle and physical factors, current smoking (NIE HR 1.05, 95% CI 1.02–1.07) and high waist-hip ratio (NIE HR 1.06, 95% CI 1.04–1.08) were the strongest mediators of the association between anxiety disorder and incident CVD, mediating 11.9% and 13.4% of the association respectively (Table 2 and Figure 2). All of the metabolic factors mediated this association with the largest contribution from systolic blood pressure (NIE HR 1.04, 95% CI 1.01–1.08) which mediated 10.4%. Similarly, current smoking (NIE HR 1.10, 95% CI 1.08–1.13) and high waist-hip ratio (NIE HR 1.08, 95% CI 1.07–1.10) were the strongest mediators of the association between depression and incident CVD, mediating 17.3% and 14.2% of the association respectively. In contrast to anxiety disorder, CRP (NIE HR 1.06, 95% CI 1.05–1.08) made the largest contribution to mediation (10.8%) among the biomarkers and systolic blood pressure made the lowest (4.3%), whose CI crossed the null value.

**Figure 2.**
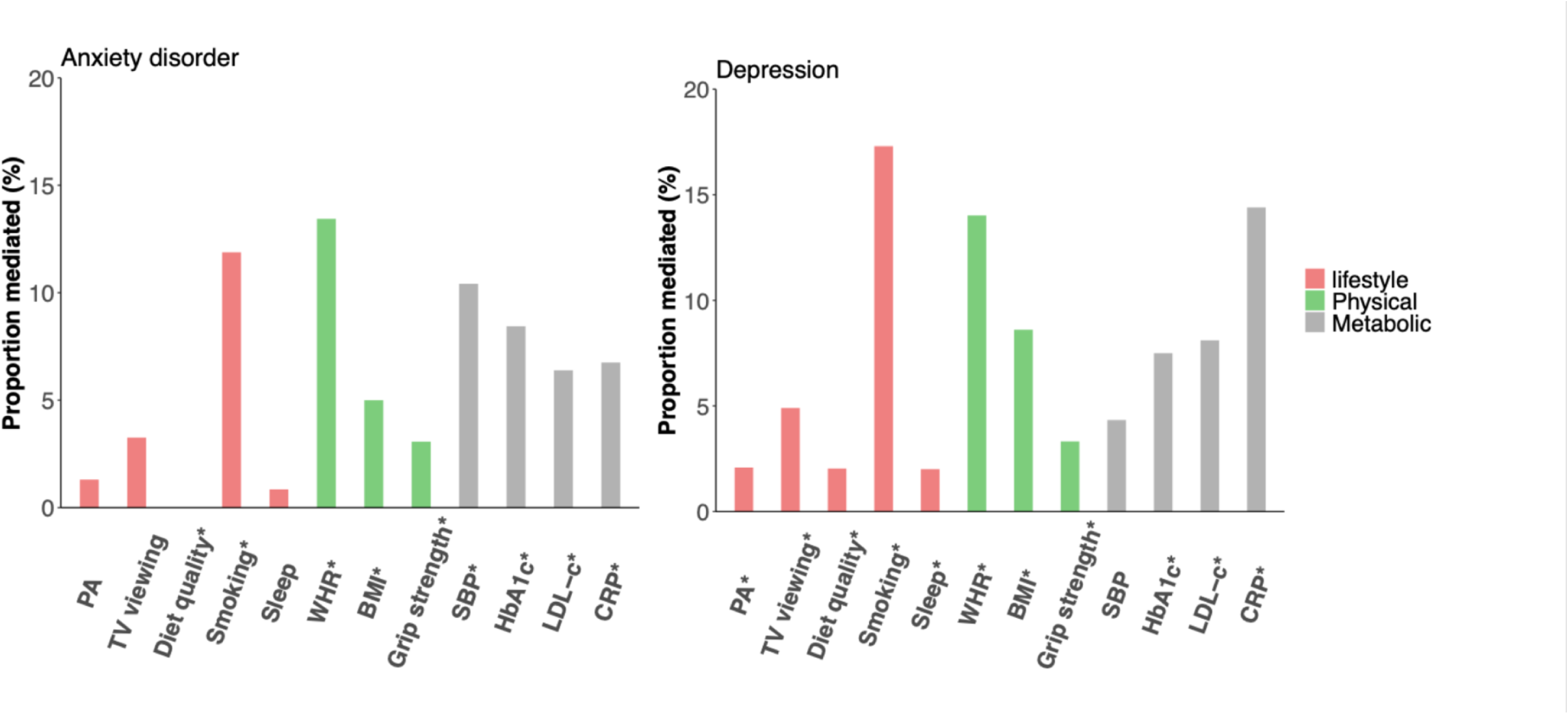
Proportions mediated by lifestyle, physical and metabolic factors of the associations between anxiety disorder and depression and incident cardiovascular disease PA, physical activity; TV, television; WHR, waist-hip ratio; SBP, systolic blood pressure; HbA1c, haemoglobin A1c; LDL-c, low-density lipoprotein cholesterol; CRP, C-reactive protein * P-value≤0.05

**Table 2.** Effect size estimates and proportions mediated via lifestyle, physical and metabolic factors of the associations between anxiety disorder and depression and incident cardiovascular disease

|  | Anxiety disorder |  |  |  |  |  | Depression |  |  |  |  |  |
| --- | --- | --- | --- | --- | --- | --- | --- | --- | --- | --- | --- | --- |
|  | Natural indirect effect |  |  | Proportion mediated |  |  | Natural indirect effect |  |  | Proportion mediated |  |  |
|  | HR | 95% CI |  | % | 95% CI |  | HR | 95% CI |  | % | 95% CI |  |
| Lifestyle factors: |  |  |  |  |  |  |  |  |  |  |  |  |
| Physical activity | 1.01 | 1.00 | 1.01 | 1.30 | -0.35 | 5.0 | 1.01 | 1.01 | 1.02 | 2.08 | 1.16 | 3.3 |
| TV viewing | 1.01 | 1.00 | 1.02 | 3.26 | -1.35 | 9.5 | 1.01 | 1.00 | 1.02 | 2.04 | 0.89 | 3.3 |
| Diet quality | 1.00 | 0.99 | 1.01 | 0.00 | 0.00 | 0.0 | 1.03 | 1.02 | 1.04 | 4.90 | 3.11 | 6.9 |
| Smoking | 1.05 | 1.02 | 1.07 | 11.88 | 4.18 | 37.8 | 1.10 | 1.08 | 1.13 | 17.30 | 13.22 | 22.7 |
| Sleep duration | 1.00 | 1.00 | 1.01 | 0.85 | -1.04 | 3.7 | 1.01 | 1.01 | 1.02 | 2.02 | 0.98 | 3.0 |
| Physical factors: |  |  |  |  |  |  |  |  |  |  |  |  |
| WHR | 1.06 | 1.04 | 1.08 | 13.44 | 7.89 | 43.0 | 1.08 | 1.07 | 1.10 | 14.02 | 10.80 | 18.7 |
| BMI | 1.02 | 1.01 | 1.03 | 5.00 | 1.33 | 18.2 | 1.05 | 1.04 | 1.06 | 8.61 | 6.25 | 12.1 |
| Grip strength | 1.01 | 1.01 | 1.02 | 3.07 | 1.04 | 10.8 | 1.02 | 1.01 | 1.03 | 3.32 | 1.96 | 4.9 |
| Metabolic factors: |  |  |  |  |  |  |  |  |  |  |  |  |
| Hypertension | 1.04 | 1.01 | 1.08 | 10.42 | 1.33 | 37.1 | 1.02 | 1.00 | 1.07 | 4.33 | -0.21 | 9.2 |
| Hyperglycaemia | 1.03 | 1.01 | 1.07 | 8.35 | 0.13 | 29.4 | 1.04 | 1.02 | 1.07 | 7.50 | 4.18 | 11.3 |
| LDL-c | 1.03 | 1.01 | 1.04 | 6.39 | 1.63 | 21.4 | 1.05 | 1.04 | 1.07 | 9.47 | 6.57 | 13.5 |
| CRP | 1.03 | 1.01 | 1.04 | 6.75 | 1.65 | 24.3 | 1.06 | 1.05 | 1.08 | 10.76 | 8.11 | 14.4 |

HR, hazard ratio; CI, confidence interval; PM, proportion mediated; NE, not estimable; TV, Television; WHR, waist and hip ratio; BMI, body mass index; SBP, systolic blood pressure; HbA1c, haemoglobin A1c; LDL-c, low-density lipoprotein cholesterol; CRP, C-reactive protein All models were adjusted for age, sex, ethnic group, and deprivation level.

The results of the sensitivity analysis are shown in Supplementary Tables 3, 4, and 5 and Supplementary Figures 2 and 3. Adding self-reported depression diagnosis showed similar trends of mediation with larger proportions mediated compared with our main findings. However, for anxiety disorder, whilst current smoking and waist-hip ratio remained the strongest mediators among lifestyle and physical factors, respectively, only LDL-c remained a significant metabolic mediator.

## Discussion

### Primary findings

To our knowledge, this is the first study to comprehensively investigate and compare the pathways linking anxiety disorder and depression to incident CVD. Whilst smoking and central obesity were common mediators of both anxiety disorder and depression, systolic blood pressure was more specific to anxiety disorder and CRP to depression.

### Comparisons with the literature

Overall, our findings are consistent with the previous findings. Previous studies have shown that anxiety disorder and depression were associated with increased risks of smoking and obesity [18–21]. Furthermore, a prospective study demonstrated that obesity mediated around 10% of the association between self-reported depression and non-fatal CVD [7]. Distinct pathways for anxiety disorder and depression have also been suggested. A meta-analysis and animal study have shown that anxiety disorder increases a risk of hypertension [22, 23], whilst a prospective study demonstrated that CRP mediated 8% of the association between depression and CVD [7]. But CRP needs to be interpreted with causation because it is a proxy marker of other causal factors rather than a causal risk factor itself [24].

The observed pathways are also theoretically and biologically plausible. Some researchers have explained the common associations of anxiety disorder and depression with smoking by applying various theoretical frameworks [25, 26]. These frameworks link latent factors behind both these mental disorders with resultant smoking behaviour. For example, low distress tolerance (behind anxiety and depression) induces smoking behaviour because people with this latent factor tend to effectively cope with stress with less effort. Additionally, the researchers have suggested the broader applicability of this framework to other unhealthy behaviours, which could also lead to obesity [25]. On the other hand, whilst anxiety disorder could rise blood pressure through persistent stress, which produces cortisol to increase vascular resistance [22, 27], depression could increase inflammation through a leaky gut, which promotes endotoxin translocation [28]. Indeed, patients with depression had higher antibodies against gut bacteria than those in comparison [29]. However, further research is required to corroborate and fully understand these pathways. Also, to explore whether and how comorbid anxiety disorder and depression interact to influence these metabolic pathways and whether the diagnoses represent different manifestations of a broader group of affective disorders, given their shared genetic risk factors.

It should also be noted that although only LDL-c remained a significant mediator for anxiety, this would not invalidate our main findings. This may indicate that, in contrast to LDL-c, hypertension is more specific to severe anxiety conditions.

### Strengths and Limitations

We established the temporal order of the exposures, mediators, and outcome by only including participants who experienced anxiety disorder or depression at least one year before the baseline and those who had no CVD history before the baseline.

There are some limitations to our study. Firstly, ascertainment of anxiety disorder and depression using hospital admission may be incomplete because it is less likely to include people with milder conditions. However, we assessed how this affected our main results through the inclusion of self-reports in the sensitivity analysis. Secondly, exposure-induced variables could confound the association between mediators and the outcome. For example, waist-hip ratio could be associated with both CRP and CVD risk. Because all mediators were measured at the same time-point we could not determine the temporal sequence nor adjust for these potential confounders. Thirdly, some mediators could be on the same pathway, so interpreting each mediated effect needs care. For example, smoking could lead to CVD through elevated systolic blood pressure. Therefore, the proportion mediated estimated in this study should not be summed. Lastly, UK Biobank participants are more likely to be white, affluent, and healthy than the national survey data, so applying our findings to the whole UK population may be limited.

### Implications

Because of the increasing number of people with anxiety disorder and depression worldwide, the prevention of CVD in this subpopulation will be of greater interest. Our study found underlying pathways of anxiety disorder and depression to CVD and would inform which risk factors to target among people with these disorders in order to reduce their higher risk of developing CVD. Whilst risk factors on the pathway have been reported to date [30], they need to be prioritised because of limited resources. More importantly, our findings raise the question as to why anxiety disorder and depression partly operate through different pathways to CVD risk and they also suggest that those with anxiety disorder or depression need different prevention strategies based on their distinct pathways.

### Conclusions

Our study examined whether lifestyle, physical and metabolic factors mediate the associations between anxiety disorder and depression and incident CVD and elucidated shared and distinct pathways. Smoking and central adiposity were the strongest mediators for both anxiety disorder and depression. Systolic blood pressure was the strong metabolic mediator of anxiety disorder whilst CRP was a major mediator of depression. Our findings inform how best to target CVD prevention strategies among people with established anxiety disorder or depression.

## Data Availability

UK Biobank data can be requested by bona fide researchers for approved projects, including replication, through https://www.ukbiobank.ac.uk/.

## Acknowledgements

We are grateful to the UK Biobank participants. This research has been conducted using the UK Biobank Resource under application number 7155. We are also grateful to the Medical Research Council and the University of Edinburgh/University of Glasgow. This work was supported by the Medical Research Council (MR/N013166/1-LGH/MS/MED2525).

## Author Contribution

SN: conceptualisation, formal analysis, writing – original draft

CM: writing – review & editing

FH, JP: conceptualisation, investigation, writing – review & editing

