## Supplementary materials for "Shared and distinct pathways from anxiety disorder and depression to cardiovascular disease: a UK Biobank prospective cohort study"

Supplementary Table 1. Associations between anxiety disorder and depression and potential mediators

Supplementary Table 2. Associations between potential mediators and incident cardiovascular disease

Supplementary Table 3. Associations between anxiety disorder and depression (ascertained through hospital admission data and self-reports) and potential mediators

Supplementary Table 4. Associations between potential mediators and incident cardiovascular disease adjusted for anxiety disorder or depression (ascertained through hospital admission data and self-reports)

Supplementary Table 5. Effect size estimates and proportions mediated via lifestyle, physical and metabolic factors of the associations between anxiety disorder and depression (ascertained through hospital admission data and self-reports) and incident cardiovascular disease

Supplementary Figure 1. Associations between anxiety disorder and depression and incident cardiovascular disease

Supplementary Figure 2. Associations between anxiety disorder and depression (ascertained through hospital admission data and self-reports) and incident cardiovascular disease

Supplementary Figure 3. Proportions mediated by lifestyle, physical and metabolic factors of the associations between anxiety disorder (ascertained through hospital admission data and self-reports) and depression and incident cardiovascular disease

Supplementary Table 1. Associations between anxiety disorder and depression and potential mediators

|  | Anxiety disorder  N = 254,695 | | Depression  N = 254,695 | |
| --- | --- | --- | --- | --- |
|  | RR | 95% CI | RR | 95% CI |
| MET-minutes/week <1000 | 1.13 | 1.00, 1.27 | 1.29 | 1.20, 1.39 |
| TV viewing ≥3 hours/day | 1.09 | 1.01, 1.18 | 1.20 | 1.14, 1.26 |
| Diet quality score <5 | 0.99 | 0.87, 1.13 | 1.18 | 1.09, 1.27 |
| Current smoking | 1.58 | 1.30, 1.92 | 2.21 | 2.00, 2.44 |
| Sleep duration <7 hours/day | 1.08 | 0.93, 1.24 | 1.27 | 1.17, 1.39 |
| Waist-hip ratio  ≥0.90 (male) ≥0.85 (female) | 1.21 | 1.13, 1.30 | 1.32 | 1.27, 1.38 |
| Body mass index ≥30kg/m2 | 1.23 | 1.07, 1.41 | 1.59 | 1.47, 1.71 |
| Grip strength  <36 Kg (male) <21 Kg (female) | 1.16 | 1.08, 1.25 | 1.25 | 1.20, 1.31 |
| SBP ≥140mmHg or medication | 1.10 | 1.01, 1.19 | 1.05 | 0.99, 1.11 |
| HbA1c ≥ 48 mmol/mol or medication | 1.69 | 1.19, 2.41 | 1.89 | 1.50, 2.37 |
| LDL-c ≥ 4.9 mmol/L or medication | 1.28 | 1.11, 1.48 | 1.60 | 1.47, 1.74 |
| CRP >3mg/L | 1.26 | 1.10, 1.45 | 1.62 | 1.50, 1.75 |

All models were adjusted for anxiety disorder or depression, age, sex, ethnic group, and deprivation level.

N, number; RR, risk ratio; CI, confidence interval; MET, metabolic equivalent; TV, television; SBP, systolic blood pressure; HbA1c, haemoglobin A1c; LDL-c, low-density lipoprotein cholesterol; CRP, C-reactive protein

Supplementary Table 2. Associations between potential mediators and incident cardiovascular disease adjusted for anxiety disorder or depression

|  | Anxiety disorder  N = 254,695 | | Depression  N = 254,695 | |
| --- | --- | --- | --- | --- |
|  | HR | 95% CI | HR | 95% CI |
| MET-minutes/week <1000 | 1.05 | 1.00, 1.09 | 1.04 | 1.00, 1.09 |
| TV viewing ≥3 hours/day | 1.10 | 1.06, 1.15 | 1.10 | 1.06, 1.15 |
| Diet quality score <5 | 1.08 | 1.03, 1.12 | 1.08 | 1.03, 1.12 |
| Current smoking | 1.81 | 1.71, 1.92 | 1.80 | 1.70, 1.91 |
| Sleep duration <7 hours/day | 1.13 | 1.08, 1.18 | 1.13 | 1.08, 1.18 |
| Waist-hip ratio  ≥0.90 (male) ≥0.85 (female) | 1.15 | 1.10, 1.21 | 1.15 | 1.10, 1.21 |
| Body mass index ≥30kg/m2 | 1.07 | 1.02, 1.12 | 1.07 | 1.02, 1.12 |
| Grip strength  <36 Kg (male) <21 Kg (female) | 1.11 | 1.06, 1.16 | 1.11 | 1.06, 1.15 |
| SBP ≥140mmHg or medication | 1.53 | 1.46, 1.61 | 1.53 | 1.47, 1.61 |
| HbA1c ≥ 48 mmol/mol or medication | 1.62 | 1.50, 1.75 | 1.62 | 1.50, 1.75 |
| LDL-c ≥ 4.9 mmol/L or medication | 1.12 | 1.07, 1.18 | 1.12 | 1.07, 1.17 |
| CRP >3mg/L | 1.25 | 1.19, 1.31 | 1.25 | 1.19, 1.30 |

The model was adjusted for anxiety disorder or depression, age, sex, ethnic group, deprivation level, and all mediators.

N, number; HR, hazard ratio; CI, confidence interval; MET, metabolic equivalent; TV, television; SBP, systolic blood pressure; HbA1c, haemoglobin A1c; LDL-c, low-density lipoprotein cholesterol; CRP, C-reactive protein

Supplementary Table 3. Associations between anxiety disorder and depression (ascertained through hospital admission data and self-reports) and potential mediators

|  | Anxiety disorder  N = 254,695 | | Depression  N = 254,695 | |
| --- | --- | --- | --- | --- |
|  | RR | 95% CI | RR | 95% CI |
| MET-minutes/week <1000 | 1.12 | 1.07, 1.18 | 1.18 | 1.15, 1.21 |
| TV viewing ≥3 hours/day | 1.10 | 1.07, 1.14 | 1.09 | 1.07, 1.11 |
| Diet quality score <5 | 1.01 | 0.96, 1.07 | 1.07 | 1.04, 1.10 |
| Current smoking | 1.34 | 1.23, 1.46 | 1.55 | 1.48, 1.63 |
| Sleep duration <7 hours/day | 1.12 | 1.06, 1.19 | 1.13 | 1.09, 1.17 |
| Waist-hip ratio  ≥0.90 (male) ≥0.85 (female) | 1.10 | 1.07, 1.14 | 1.16 | 1.14, 1.18 |
| Body mass index ≥30kg/m2 | 1.06 | 1.00, 1.13 | 1.31 | 1.27, 1.36 |
| Grip strength  <36 Kg (male) <21 Kg (female) | 1.12 | 1.09, 1.16 | 1.12 | 1.10, 1.14 |
| SBP ≥140mmHg or medication | 1.02 | 0.99, 1.05 | 0.97 | 0.95, 0.99 |
| HbA1c ≥ 48 mmol/mol or medication | 1.04 | 0.87, 1.25 | 1.42 | 1.28, 1.57 |
| LDL-c ≥ 4.9 mmol/L or medication | 1.20 | 1.13, 1.28 | 1.28 | 1.24, 1.33 |
| CRP >3mg/L | 1.10 | 1.03, 1.17 | 1.28 | 1.24, 1.33 |

All models were adjusted for anxiety disorder or depression (ascertained through hospital admission data and self-reports), age, sex, ethnic group, and deprivation level.

N, number; RR, risk ratio; CI, confidence interval; MET, metabolic equivalent; TV, television; SBP, systolic blood pressure; HbA1c, haemoglobin A1c; LDL-c, low-density lipoprotein cholesterol; CRP, C-reactive protein

Supplementary Table 4. Associations between potential mediators and incident cardiovascular disease adjusted for anxiety disorder or depression (ascertained through hospital admission data and self-reports)

|  | Anxiety disorder  N = 254,695 | | Depression  N = 254,695 | |
| --- | --- | --- | --- | --- |
|  | HR | 95% CI | HR | 95% CI |
| MET-minutes/week <1000 | 1.05 | 1.00, 1.09 | 1.05 | 1.00, 1.09 |
| TV viewing ≥3 hours/day | 1.10 | 1.06, 1.15 | 1.10 | 1.06, 1.15 |
| Diet quality score <5 | 1.08 | 1.03, 1.12 | 1.08 | 1.03, 1.12 |
| Current smoking | 1.81 | 1.71, 1.92 | 1.81 | 1.71, 1.92 |
| Sleep duration <7 hours/day | 1.13 | 1.08, 1.18 | 1.13 | 1.08, 1.18 |
| Waist-hip ratio  ≥0.90 (male) ≥0.85 (female) | 1.15 | 1.10, 1.21 | 1.15 | 1.10, 1.21 |
| Body mass index ≥30kg/m2 | 1.07 | 1.02, 1.12 | 1.07 | 1.02, 1.12 |
| Grip strength  <36 Kg (male) <21 Kg (female) | 1.11 | 1.06, 1.16 | 1.11 | 1.06, 1.16 |
| SBP ≥140mmHg or medication | 1.53 | 1.46, 1.61 | 1.53 | 1.46, 1.61 |
| HbA1c ≥ 48 mmol/mol or medication | 1.62 | 1.50, 1.75 | 1.62 | 1.50, 1.75 |
| LDL-c ≥ 4.9 mmol/L or medication | 1.12 | 1.07, 1.18 | 1.12 | 1.07, 1.18 |
| CRP >3mg/L | 1.25 | 1.19, 1.31 | 1.25 | 1.19, 1.31 |

The model was adjusted for anxiety disorder or depression (ascertained through hospital admission data and self-reports), age, sex, ethnic group, deprivation level, and all mediators.

N, number; HR, hazard ratio; CI, confidence interval; MET, metabolic equivalent; TV, television; SBP, systolic blood pressure; HbA1c, haemoglobin A1c; LDL-c, low-density lipoprotein cholesterol; CRP, C-reactive protein

Supplementary Table 5 Effect size estimates and proportions mediated via lifestyle, physical and metabolic factors of the associations between anxiety disorder and depression (ascertained through hospital admission data and self-reports) and incident cardiovascular disease

|  | Anxiety disorder | | | | | | Depression | | | | | |
| --- | --- | --- | --- | --- | --- | --- | --- | --- | --- | --- | --- | --- |
|  | Natural indirect effect | | | Proportion mediated | | | Natural indirect effect | | | Proportion mediated | | |
|  | HR | 95% CI | | % | 95% CI | | HR | 95% CI | | % | 95% CI | |
| Lifestyle factors: |  |  |  |  |  |  |  |  |  |  |  |  |
| Physical activity | 1.00 | 1.00 | 1.01 | 2.88 | 0.97 | 11.5 | 1.01 | 1.00 | 1.01 | 5.13 | 2.02 | 18.3 |
| TV viewing | 1.01 | 1.01 | 1.02 | 8.27 | 2.30 | 26.5 | 1.00 | 1.00 | 1.01 | 3.03 | 1.07 | 13.2 |
| Diet quality | 1.00 | 1.00 | 1.00 | 0.53 | -2.93 | 5.7 | 1.01 | 1.01 | 1.02 | 8.98 | 4.30 | 32.7 |
| Smoking | 1.03 | 1.02 | 1.04 | 16.07 | 5.23 | 56.6 | 1.05 | 1.04 | 1.05 | 31.06 | 18.00 | 110.5 |
| Sleep duration | 1.01 | 1.00 | 1.01 | 2.96 | 0.46 | 11.4 | 1.01 | 1.00 | 1.01 | 3.92 | 1.66 | 14.4 |
| Physical factors: |  |  |  |  |  |  |  |  |  |  |  |  |
| WHR | 1.03 | 1.02 | 1.05 | 14.85 | 7.08 | 47.7 | 1.04 | 1.03 | 1.05 | 26.70 | 15.16 | 75.8 |
| BMI | 1.01 | 1.00 | 1.01 | 3.21 | -1.77 | 11.0 | 1.03 | 1.02 | 1.03 | 18.09 | 10.14 | 55.0 |
| Grip strength | 1.01 | 0.00 | 1.01 | 5.23 | 2.03 | 21.0 | 1.01 | 1.01 | 1.01 | 6.50 | 3.05 | 21.3 |
| Metabolic factors: |  |  |  |  |  |  |  |  |  |  |  |  |
| Hypertension | 1.01 | 1.00 | 1.02 | 5.91 | -6.23 | 23.1 | 0.99 | 0.98 | 1.00 | 0.00 | 0.00 | 0.0 |
| Hyperglycaemia | 1.00 | 0.99 | 1.01 | 1.47 | -8.82 | 11.9 | 1.02 | 1.01 | 1.03 | 13.88 | 6.28 | 47.3 |
| LDL-c | 1.02 | 1.01 | 1.03 | 10.51 | 3.76 | 37.2 | 1.03 | 1.02 | 1.03 | 17.42 | 9.34 | 54.3 |
| CRP | 1.01 | 1.00 | 1.02 | 6.08 | -1.20 | 24.3 | 1.03 | 1.02 | 1.03 | 19.24 | 10.64 | 65.7 |

HR, hazard ratio; CI, confidence interval; N, number; NE, not estimable; TV, television; WHR, waist and hip ratio; BMI, body mass index; SBP, systolic blood pressure; HbA1c, haemoglobin A1c; LDL-c, low-density lipoprotein cholesterol; CRP, C-reactive protein

All models were adjusted for age, sex, ethnic group, and deprivation level.

Supplementary Figure 1. Associations between anxiety disorder and depression and incident cardiovascular disease

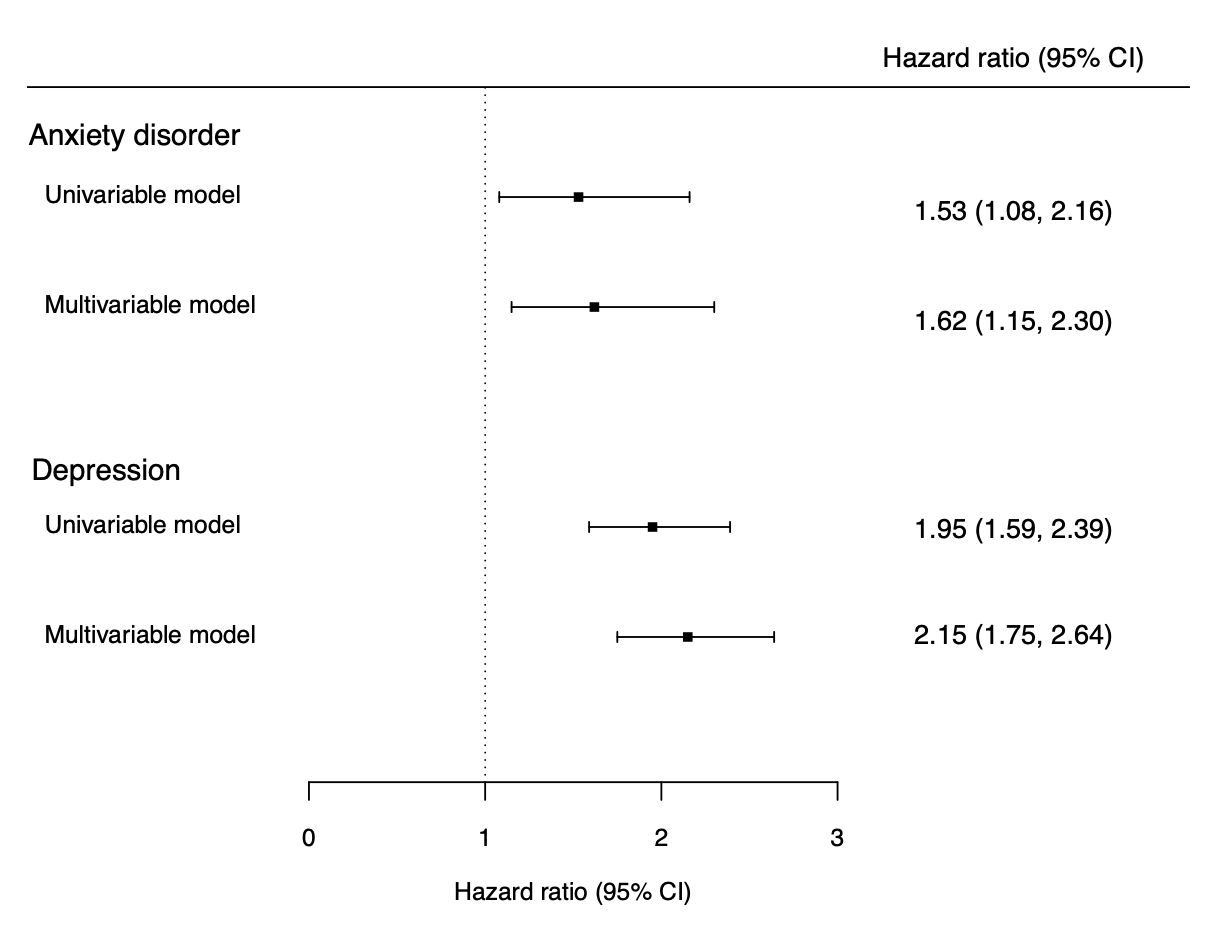

Multivariable models included age, sex, ethnic group, and deprivation level.

CI, confidence interval

Supplementary Figure 2. Associations between anxiety disorder and depression (ascertained through hospital admission data and self-reports) and incident cardiovascular disease

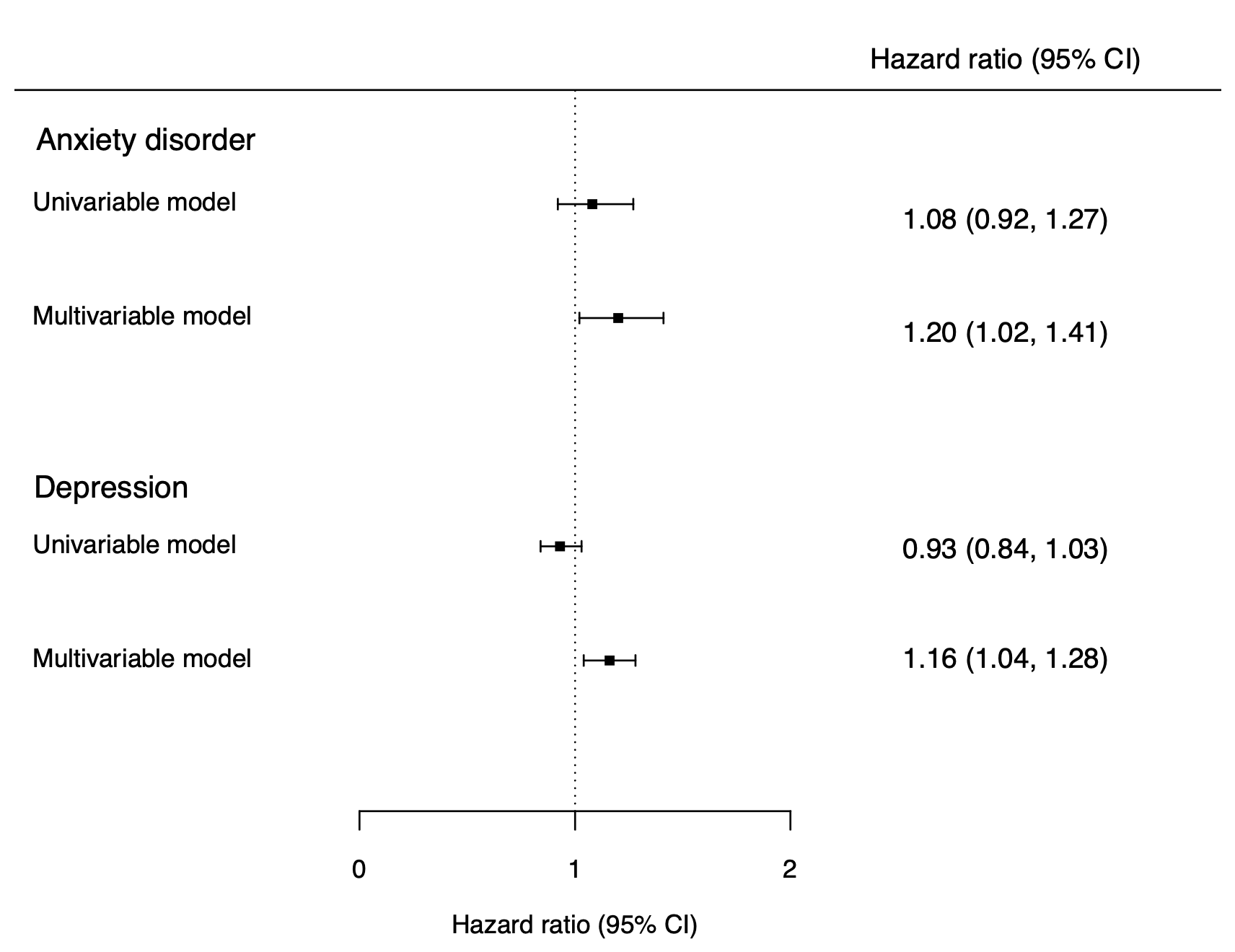

Multivariable models included age, sex, ethnic group, and deprivation level.

CI, confidence interval

Supplementary Figure 3. Proportions mediated by lifestyle, physical and metabolic factors of the associations between anxiety disorder (ascertained through hospital admission data and self-reports) and depression and incident cardiovascular disease

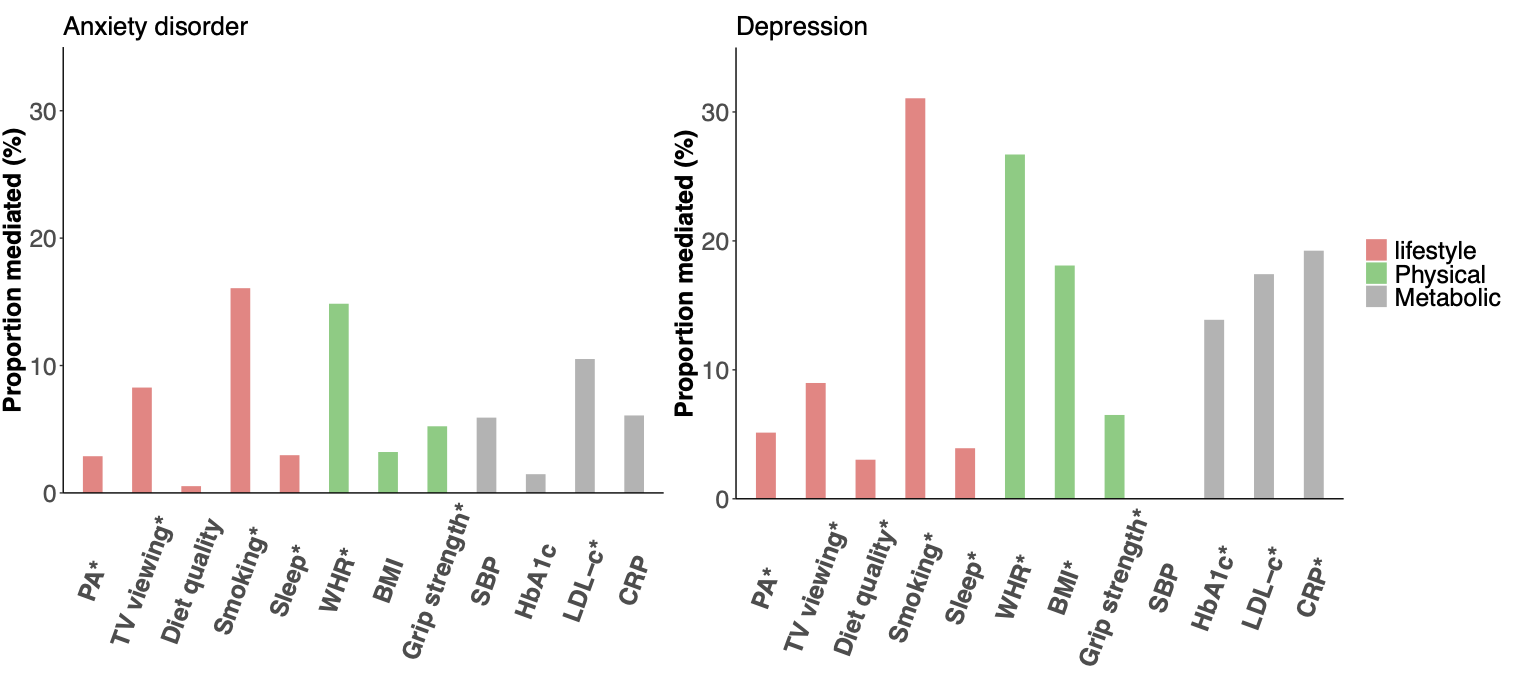

PA, physical activity; TV, television; WHR, waist-hip ratio; SBP, systolic blood pressure; HbA1c, haemoglobin A1c; LDL-c, low-density lipoprotein cholesterol; CRP, C-reactive protein

* P-value≤0.05
